# Global Parkinson’s Genetics Program (GP2) Monogenic Network Protocol: Elucidating causative gene variants in hereditary Parkinson’s disease

**DOI:** 10.1101/2022.12.01.22282794

**Authors:** Lara M. Lange, Micol Avenali, Melina Ellis, Anastasia Illarionova, Ignacio J. Keller Sarmiento, Ai-Huey Tan, Harutyun Madoev, Caterina Galandra, Johanna Junker, Karisha Roopnarain, Justin Solle, Claire Wegel, Zih-Hua Fang, Peter Heutink, Kishore R. Kumar, Shen-Yang Lim, Enza Maria Valente, Mike Nalls, Cornelis Blauwendraat, Andrew Singleton, Niccolo Mencacci, Katja Lohmann, Christine Klein, the Global Parkinson’s Genetic Program (GP2)

## Abstract

The Monogenic Network of the Global Parkinson’s Genetics Program (GP2) aims to create an efficient infrastructure to accelerate the identification of novel genetic causes of Parkinson’s disease (PD) and to improve our understanding of already identified genetic causes, such as reduced penetrance and variable clinical expressivity of known disease-causing variants. We aim to perform short- and long-read whole-genome sequencing for up to 10,000 patients with parkinsonism.

## Main text

The Global Parkinson’s Genetics Program (GP2, http://gp2.org/) is an international collaborative effort that aims to substantially improve our understanding of the role that genetics plays in Parkinson’s disease (PD) and to make this knowledge globally available and actionable.^1^ The Monogenic Network of GP2 creates an efficient infrastructure to accelerate the identification of novel genetic causes of parkinsonism but also to improve our current understanding of known genetic causes, such as reduced penetrance and variable clinical expressivity of known genetic variants. To address a large gap in our knowledge of PD due to the lack of genetic studies in diverse populations, we bring together clinicians, researchers and existing PD consortia from around the globe with a particular emphasis on historically underrepresented populations. We collect and generate clinical and genetic data of PD patients and families, harmonize and democratize data as well as the access to it, and develop analytical resources. All clinicians and researchers interested in collaborative PD genetics research are welcome to be part of the GP2 Monogenic Network and share data and biomaterials of PD patients and family members with i) a known monogenic cause of disease or ii) an unknown but suspected monogenic cause, e.g., based on a young age at onset (AAO) or a positive family history.

The workflow of the Monogenic Network of GP2 is shown in Figure 1. The first step is to register to the Monogenic Portal (https://monogenic.gp2.org/monogenicportal.html). This online platform serves as a secure bidirectional site to upload and download clinical and genetic data regarding patients/families with potentially monogenic PD. More specifically, the Portal allows participants to readily access information on the project, facilitates the collection of detailed data, and enables a deeper analysis of genetic, clinical-demographic, and environmental factors influencing PD development and expression.^2,3^ Key items include a registration site to submit information regarding institutional ethical clearance for international data and sample sharing and an electronic case report form (eCRF) to submit pseudonymized data of patients/families securely. As a prerequisite, every research institution has to obtain institutional ethics approval. Researchers are required to submit their ethics documents through the Portal for GP2 compliance review. Following compliance approval, research agreements will be initiated before data and samples can be shared.

**Figure 1.**
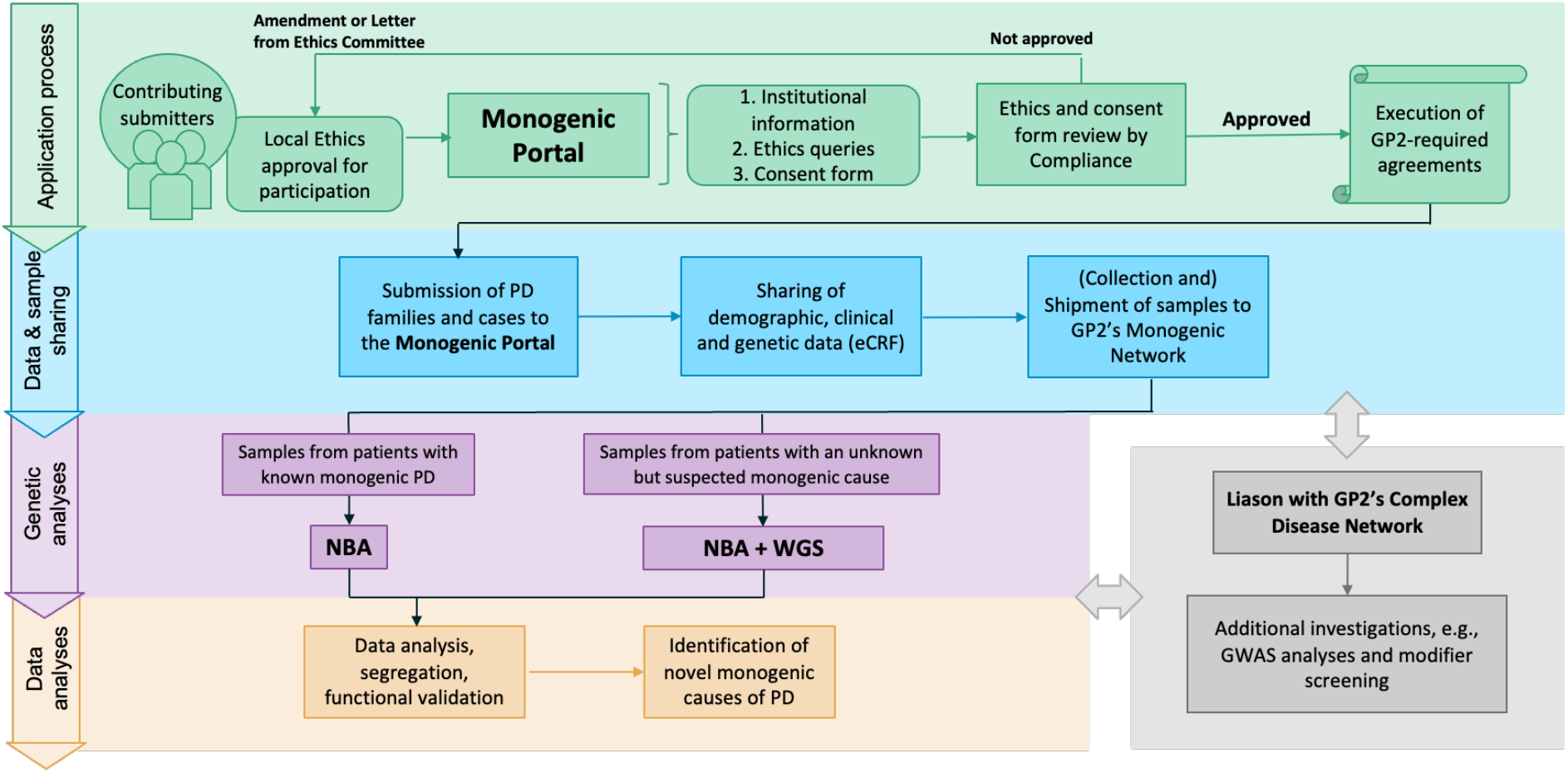
Workflow of the GP2 Monogenic Network. eCRF = electronic case report form, GP2 = Global Parkinson’s Genetics Program, GWAS = genome-wide association studies, NBA = NeuroBooster Array, PD = 8abelled8’s disease, WGS = Whole-genome sequencing There are four steps: I) The application process, where interested collaborators register to the Monogenic Portal, ethics documents are reviewed, and the required paperwork is executed. II) The data and sample sharing, where collaborators ship their samples to GP2’s Monogenic Network and share the respective clinical data for these patients by filling out the eCRFs in the Portal. III) The genetic analyses that are performed by GP2’s Monogenic Network, in particular, NBA genotyping for all patients and both NBA and WGS for a subset of unsolved, apparently monogenic cases. IV) The data analyses where generated data are interpreted, and results validated.

The eCRF (https://monogenic.gp2.org/testing/ecrf1) consists of multiple questionnaires focusing on i) demographics and basic clinical details, ii) family history, iii) PD clinical features, investigations, and treatments, and iv) relevant environmental or acquired factors prior to PD motor symptom onset. Within the Portal, collaborators also gain access to research-based genetic results regarding their own cases and monitor the genetic analysis process.

All samples submitted to the Monogenic Network undergo quality control and are adequately prepared for genotyping and sequencing. The preferred sample type is high-quality genomic DNA (gDNA) obtained from blood or alternatively saliva, but fresh blood in an EDTA tube (10ml), and frozen EDTA blood or saliva can also be accepted.

The Monogenic Network of GP2 focuses on monogenic causes of the disease, and aims to identify and collect cases with a higher probability of finding novel PD-causing genes. Priority is given to families with a greater number of available samples, given that this is likely to improve the filtering procedure of thousands of genetic variants from WGS data. We also prioritize consanguineous families which have a higher chance of carrying homozygous or compound-heterozygous recessive genetic variants. Also, a younger AAO of PD is prioritized given the increased genetic load in early-onset PD.^4^ Furthermore, we preferentially include samples from underrepresented populations to support our goal of greater representation of these groups.^5^ Additionally, the Monogenic Network is also interested in patients and families with genetic variants in already known PD genes.

Regardless of whether samples have been genetically tested before and whether they are already known to carry PD-related mutations, every sample will undergo genotyping with the NeuroBooster Array (NBA). This array includes 1.9 million markers from the Illumina Global Diversity Array and more than 95,000 neurological disease-oriented and population-specific variants, including several hundred known mutations in PD-related genes (https://github.com/GP2code/Neuro_Booster_Array). We will also test for large deletions and duplications encompassing known PD genes, which represent another frequent cause of monogenic PD.^6,7^ Second, about 10,000 prioritized cases negative for mutations in known genes will undergo Illumina short-read WGS. We use the functional equivalence pipeline^8^ implemented at the Broad Institute to produce alignments and small variant calls against the GRCh38DH reference genome, as well as the Broad Institute’s joint discovery pipeline to produce a set of joint-genotyped variants for all the samples that pass WGS quality controls following the quality metrics defined by the Accelerating Medicines Partnership Parkinson’s Disease program (AMP-PD; https://amp-pd.org).9 After joint-genotyping, we retain high-quality variants that are flagged as PASS after variant quality score recalibration, with a call rate >0.95, genotype quality >20, read depth >5, and heterozygous allele balance between 0.25 and 0.75. Additionally, we discover large structural variants (>50 base pairs) for each sample using Parliament2 pipeline^10^ and perform joint-genotyping using graphtyper2^11^. Following variant quality control, we annotate variants with Ensembl Variant Effect Predictor^12^ to prioritize candidate single-nucleotide variants, indels, and structural variants based on population frequencies, segregation, and *in silico* predictor scores. All the pipelines and scripts used for monogenic data analyses are available via GitHub (https://github.com/GP2code/GP2-WorkingGroups/tree/main/MN-DAWG-Monogenic-Data-Analysis). In a third step, a subset of prioritized unsolved cases (n = 1,000) will undergo long-read WGS with Oxford Nanopore technologies which will be used to generate population-specific genome assembly, haplotype phasing, and the detection of repeat expansions and structural variants. In addition, we will integrate the resulting genetic data with the clinical information available from the Portal (e.g., AAO and clinical features).

To date, the Monogenic Network has contacted ∼250 potential contributors from >60 different countries (Figure 2). We are about to complete a 500-short-read WGS pilot project including 16 research teams from 10 countries covering five continents. Around three quarters of this pilot cohort were familial cases, whereas the remaining patients were singleton cases with an early age at onset of disease (≤40 years); twelve of these singletons were included as parent-offspring trios (index patient plus both clinically unaffected parents). Notably, ∼20% of selected patients came from underrepresented populations, mainly South America and South-East Asia. Following the pilot, another ∼2000 samples have already been submitted to the Monogenic Network and are currently undergoing genotyping and sequencing.

**Figure 2.**
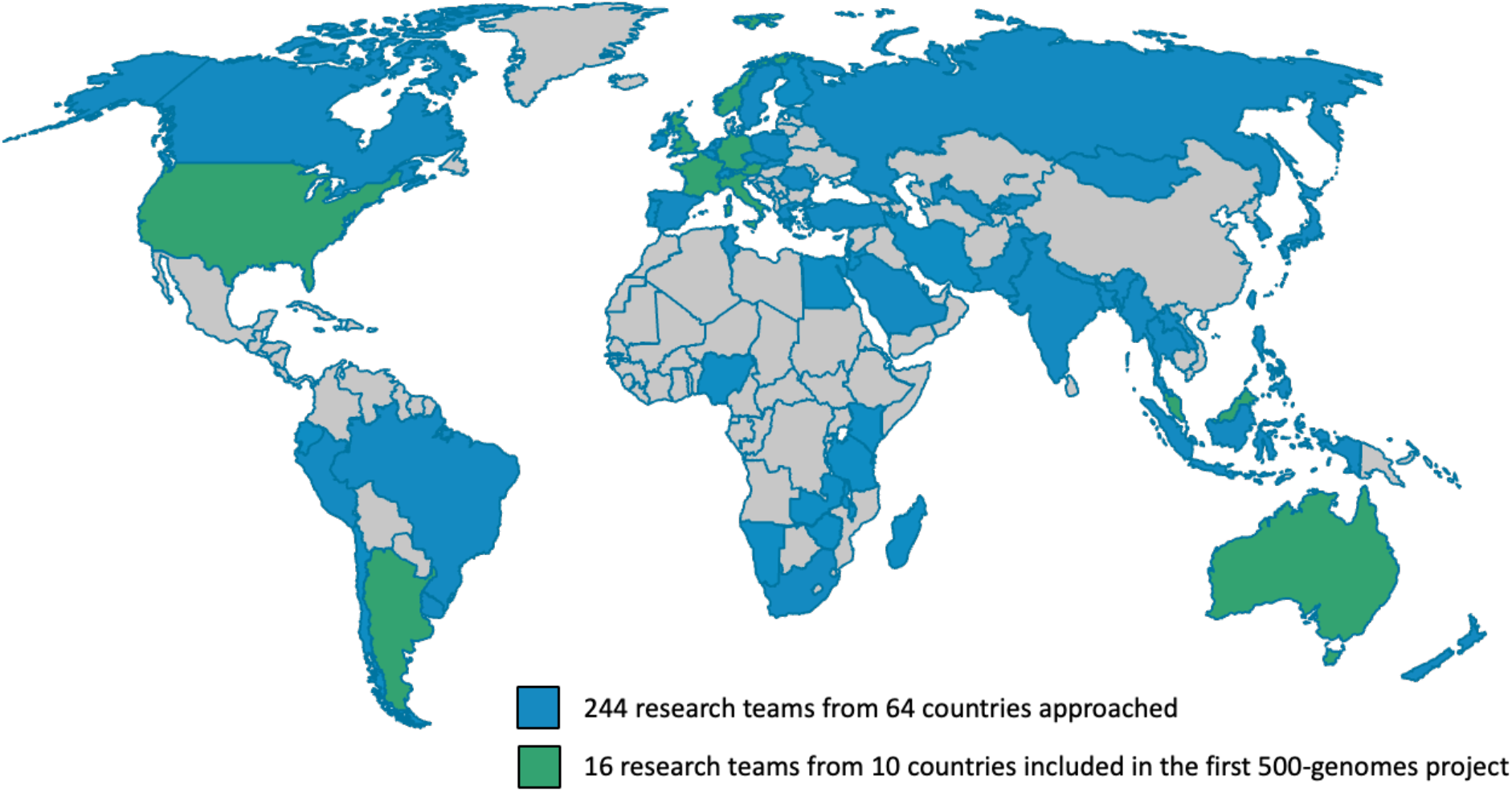
Outreach of the Monogenic Network (world map) and selected cohort for the 500-genomes pilot project (September 2022). Highlighted in blue are countries where potential collaborators have been contacted, and highlighted in green are the countries from which research teams have been involved in the 500-genomes pilot project. Over 60 of these teams from almost 40 countries are already participating in the GP2 project.

Hurdles encountered during the project so far include the administrative burden of ensuring compliance with ethics and local and international regulatory requirements. Furthermore, the SARS-CoV-2 pandemic has posed unique challenges by limiting personal visits and slowing recruitment. Moreover, there is a strong focus on recruiting and supporting participants from low-middle income countries that often have only limited experience, resources, and infrastructure for sample collection and processing.

The identification of genes causing monogenic PD has provided critical insights into the underlying disease pathophysiology. However, despite extensive investigations, only a minority (10-40%^13^) of likely monogenic cases receive a genetic diagnosis to date; it is, thus, imperative to investigate these unsolved families for novel PD genes. Moreover, even genetically proven cases entail significant challenges in terms of understanding the genetic modifiers and possible other additional factors underlying the variable penetrance and expressivity of many genetic mutations (e.g., *LRRK2* p.G2019S or *GBA* variants). Understanding these modifying factors is likely to provide insights into disease pathogenesis and the potential routes to new treatment. Within the Monogenic Network of GP2, we have established a workflow that allows us to work together with clinicians and researchers from around the globe, overcome national and international privacy challenges, as well as obtain DNA samples and clinical information from likely and known monogenic PD cases, and perform high throughput genetic analysis. Given the extensive infrastructure and global outreach, this project is expected to contribute substantially to our understanding of the genetic basis of monogenic PD, with a particular focus on populations that have not yet been largely addressed by existing research.

## Supporting information

Banner Authorship List

## Data Availability

GP2 has partnered with the Accelerating Medicines Partnership - Parkinson's Disease (AMP-PD; https://amp-pd.org) to share data generated by GP2. The first GP2 data was released to the AMP-PD platform in December 2021, and there will be data releases at regular intervals as the project continues.

## Acknowledgment

This research is supported by the Aligning Science Across Parkinson’s Initiative and the Michael J. Fox Foundation for Parkinson’s Research.

Data used in the preparation of this article were obtained from Global Parkinson’s Genetics Program (GP2). GP2 is funded by the Aligning Science Against Parkinson’s (ASAP) initiative and implemented by The Michael J. Fox Foundation for Parkinson’s Research (https://gp2.org). For a complete list of GP2 members see https://gp2.org.

## Author contributions

L. M. Lange, M. Avenali, M. Ellis, Z.-H. Fang, C. Galandra, P. Heutink, A. Illarionova, J. Junker, I. J. Keller Sarmiento, K. R. Kumar, S.-Y. Lim, K. Lohmann, H. Madoev, N. Mencacci, K. Roopnarain, A.-H. Tan, E. M. Valente, and C. Klein are members of the GP2 Monogenic Network, of which C. Klein is the lead, and K. Lohmann and N. Mencacci are the co-leads.

L. M. Lange was the primary contributor in the drafting of this manuscript, assisted by M. Avenali, M. Ellis, K. R. Kumar, and A.-H. Tan. Z.-H. Fang is the lead of the data analysis working group and drafted the data analysis section of this manuscript together with I. J. Keller Sarmiento, the co-lead of the data analysis working group. The manuscript draft was reviewed by all members of the Monogenic Network prior to circulation to the other co-authors. J. Solle and C. Wegel are members of GP2’s Compliance Working Group, of which J. Solle is the co-lead.

M. Nalls is the lead of the GP2 Complex Disease Data Analysis Working Group (DAWG) and assisted with developing the data analysis workflows.

A. Singleton and C. Blauwendraat are the leads of the Global Parkinson’s Genetics Program. All listed authors reviewed the manuscript and provided comments and revisions prior to submission.

## Competing Interests statement

C. Klein has received grant support from the Michael J. Fox Foundation for Parkinson’s Research and the Aligning Science Across Parkinson’s Initiative. She serves as a medical advisor to Centogene and Retromer Therapeutics and has received a speaker’s honorarium from Desitin Pharma.

S.-Y. Lim received grants from the Michael J. Fox Foundation and the Malaysian Ministry of Education Fundamental Research Grant Scheme.

A. Singleton and C. Blauwendraat are supported by the Intramural Research Program of the National Institute on Aging and have received grant support from the Michael J. Fox Foundation for Parkinson’s Research and the Aligning Science Across Parkinson’s Initiative. A. Singleton has received royalty payments related to a diagnostic for stroke.

M. Nalls is a consultant employed by Data Tecnica International. Data Tecnica is engaged in a consulting agreement with the US National Institutes of Health.

## Notes

### Author Declarations

Every single Ethics committee/IRB of every center participating in this study gave ethical approval for this work.

