## Supplementary Material for "Global Parkinson’s Genetics Program (GP2) Monogenic Network Protocol: Elucidating causative gene variants in hereditary Parkinson’s disease"

#### I. Additional methods

#### Cohort Recruitment

Researchers and clinicians have been contacted through personalized invitation emails or personal contacts. Interested PD clinicians and researchers will be initiated via introductory video conference calls covering the nature of GP2 and its goals, benefits of participation, sample and clinical data requirements, and ethical and compliance issues. If required, templates for research protocols, patient information, and consent forms are shared as needed, and assistance is provided for contributors who require help in their ethics application process.

Detailed information can also be found on the GP2 (https://gp2.org) and the Monogenic Network (https://monogenic.gp2.org/index.html) website.

#### Sample requirements and preparation

#### The Monogenic Network accepts three different sample types:

1. The preferred sample type is **high-quality genomic DNA** from blood (alternatively from saliva)
    - ✓ Required volume: >60µl
    - ✓ Required concentration: >50 ng/µl (in TE buffer)
    - ✓ Quality: OD 260/280nm: ~1.8

   *Shipping Instructions:* Submit sample in a clearly 1abelled 1.5~2.0ml microcentrifuge tube sealed with parafilm tightly. Place sealed microcentrifuge tubes in a 50ml disposable screw cap tube or a small solid box for additional insulation during shipment.

2. Alternatively, **fresh ~10 ml EDTA blood**.
   *Collecting and shipping instructions:* The tubes should be filled properly, inverted (not shaken) 10 times carefully. After collection, the tubes should be kept at 4 degrees Celsius (do not freeze) and sent off as soon as possible. Place the EDTA tube in a 50ml disposable screw cap tube or a small solid box for additional insulation during shipment. To prevent the EDTA tube from moving during shipment, fill any remaining space in the 50ml tube or box with clean tissue paper or bubble wrap before sealing.

3. If you neither have isolated DNA nor can freshly collect a sample, we can also accept (although not preferred) **frozen EDTA blood** (~5 ml) or a **frozen saliva sample**. In this case, the blood or saliva has to be shipped on dry ice.
   *Shipping Instructions:* Place the frozen EDTA/saliva tube in a 50ml precooled, disposable screw cap tube or a small solid box for additional insulation during shipment.

Every DNA sample that is shared with the Monogenic Hub undergoes quality control. If samples do not meet the criteria, they are adjusted accordingly by either sample dilution with high-performance liquid chromatography (HPLC) water or by sample concentration by evaporation using a thermoshaker at 55°C. Quality is checked with a spectrophotometer (Nanodrop™1000 Spectrophotometer). When whole blood or saliva is submitted, DNA extraction is performed following standard techniques. For DNA extraction from whole blood (fresh or frozen), the Roche High Pure Viral Nucleic Acid Large Volume Kit is used, and for frozen saliva, the Norgen Saliva DNA Isolation Kit is used. In addition, an extraction machine, AGFSTAR (AutoGen), is used for large amounts of either fresh or frozen blood, together with the FlexiGene DNA extraction Kit (Qiagen). After sample preparation, DNA samples are pipetted into tubes and sent for NBA genotyping and/or WGS.

### II. Additional Tables and Figures

**Supplementary Table 1. Sample prioritization criteria.**

| | Family history/ Available samples | Age at onset (AAO) | Ethnicity |
|---|---|---|---|
| **Very strong** | > 2 affected | <20 years | Both parents with non-European ancestry (underrepresented)* |
| **Strong** | 1 affected, both healthy parents | 21-40 years | One side/parent with non-European ancestry (underrepresented)* |
| **Medium** | 2 affected | 41-50 years | Other non-European |
| **Possibly** | 1 affected but strong positive family history | >50 years | White/European |

*: Africa, Latin America/Caribbean, Indigenous Americans/Oceania, Middle East, and non-East Asians

This table lists the criteria the Monogenic Network uses to evaluate the likelihood of a monogenic cause of the disease. Based on these criteria listed in the table as well as the presence of consanguinity, submitted samples will be prioritized for WGS.

**Supplementary Figure 1. The data generation and processing workflow for GP2 monogenic samples.**

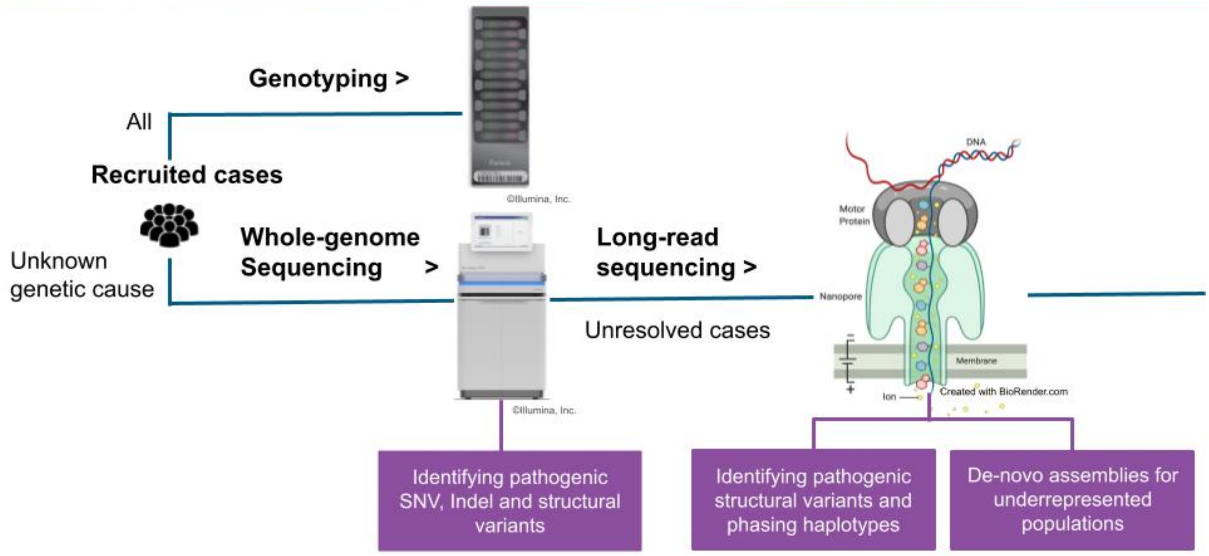
